# Sex-, and neurodegeneration-dependent effect modification in the association between thyroid function and cognitive impairment in non-depressed, non-demented elderly

**DOI:** 10.1101/2024.07.04.24309827

**Authors:** Asma Hallab, the Alzheimer’s Disease Neuroimaging Initiative

## Abstract

**Introduction:** Understanding the particularities of thyroid-cognition interactions in the elderly is crucial in assessing the risks and evaluating therapeutic options.

**Methods:** Cross-sectional analyses where participants from Alzheimer’s Disease Neuroimaging Initiative (ADNI) with mild cognitive impairment (MCI) and healthy controls (HC), with complete neurocognitive tests, thyroid stimulating hormone (TSH) <10 µIU/mL, and geriatric depression scale (GDS) <5 were eligible. Linear and logistic regression models, including testing for non-linearity, were performed. Sex and neurodegeneration-related stratifications were explored.

**Results:** Of the total 1845 participants, with a median age of 73 (IQR: 68, 78); 887 (48%) were females, and 1056 (57%) had MCI. The median TSH level was 1.70 µIU/mL (IQR: 1.15, 2.40). There was a significant association between TSH and cognition only in males (adj. *ß*_Males_: -0.40; 95% *CI*: -0.74, -0.07; *p*-value: 0.019). The odds of being diagnosed with MCI at baseline decreased with higher TSH levels in the total study population (adj. OR_Total_: 0.87; 95% *CI*: 0.79, 0.95; *p*-value: 0.002), and in males (adj. OR_Males_: 0.80; 95% *CI*: 0.70, 0.92; *p*-value: 0.001). The median TSH value was a significant cutoff in this association.

**Conclusions:** The association between thyroid function and cognitive decline in the elderly is subject to a sex-driven effect modification and depends on a cutoff value.

**Plain English summary:** The thyroid–brain association starts at very early stages of the nervous system development and plays a central role in cognition. During the aging process, the thyroid maintains an important role in modulating mental health well-being and associated risks. Older persons are at higher risk of hypothyroidism (lower functioning of thyroid hormone), which is a risk factor for reversible cognitive impairment and dementia. The current study explored the association between thyroid stimulating hormone, a central biomarker of thyroid function, and cognitive function in the elderly. People with dementia, depression, and overt hypothyroidism were excluded to better assess the risks beyond those well-established risk factors. Using different advanced statistical methods, a significant association between thyroid function and cognitive impairment was observed only in males but not females. The association was particularly relevant in older males with lower TSH levels under the median TSH value. Sex-related mechanisms and the reversibility of the association after appropriate intervention are still unclear. It is therefore important to explore thyroid-brain interactions in males and females separately and use methods testing for non-linear associations. The study design based on a cross-sectional analysis of baseline data does not imply causation and randomized longitudinal studies are needed.

**Highlights:**

- ADAS_13_ total score was negatively correlated with TSH levels in a statistically significant manner only in males.
- Higher TSH levels predicted significantly lower ADAS_13_ scores only in males.
- Lower TSH levels were significantly associated with higher odds of mild cognitive impairment only in males.
- The median TSH value was a significant cutoff point in the association between thyroid function and mild cognitive impairment.

**Graphical abstract:** 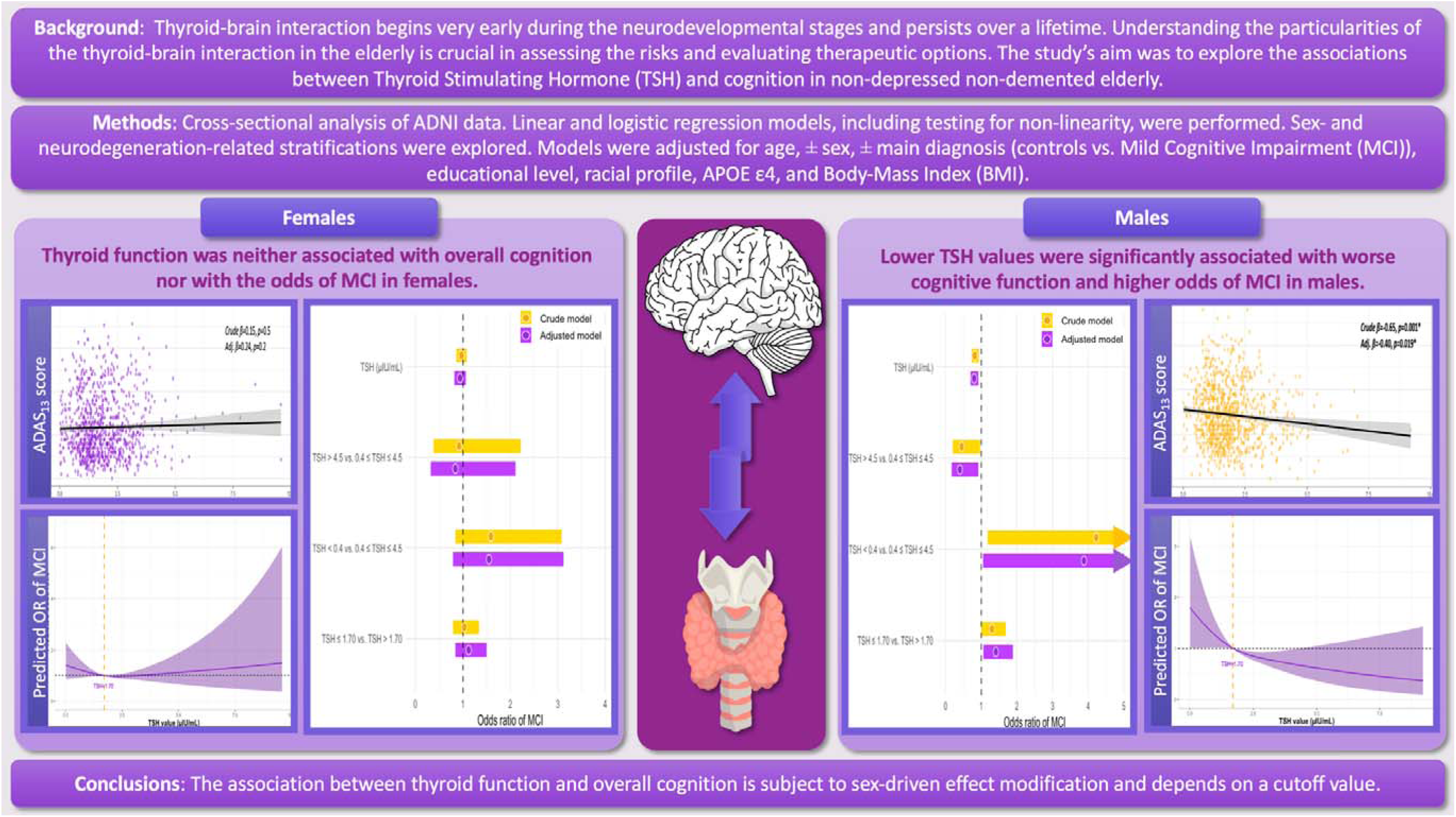

## 1. Introduction

Cognitive impairment is a serious public health concern, affecting millions worldwide and severely impacting their quality of life and independence. It is estimated that 152.8 million persons worldwide will experience some form of dementia by 2050. (1) Understanding related risk factors is crucial for preventive and interventional strategies. Various physiological systems are involved in cerebral functioning and cognition, and the thyroid plays a pivotal role in this body-brain association. (2) Thyroid-brain interactions begin very early during the neurodevelopmental stages and persist over a lifetime. (3, 4) During adulthood and the aging process, thyroid hormones maintain a central role in modulating the risk of mental health, being significantly associated, in a sex-dependent manner, (5) with the onset and complications of affective and psychotic disorders. (6–11) Thus, the impact of thyroid hormones on cognitive functions, particularly the association between dysthyroidism, whether hypothyroidism (low functioning) or hyperthyroidism (high functioning) on one side and cognitive decline during the aging process on the other side, is an emerging research field with multiple edges and of high complexity. Studies in adults and the elderly reported controversial outcomes, depending on the underlying demographical particularities and the assessed physiological risk factors. (12–16) Owing to a physiologically increased risk of subclinical hypothyroidism in the elderly, (17, 18) concomitantly with an increased risk of neurodegeneration and cognitive decline, understanding the interaction between both conditions is crucial to minimize the risks. Furthermore, women are more prone to thyroid disorders (19) and exhibit different patterns of cognitive decline than men. (20, 21) It is therefore unclear how significant sex-related biological factors might interfere with the thyroid-cognition association.

The general aim of this study was to explore the associations between thyroid function and cognition in non-depressed non-demented elderly and identify the major effect modifiers. The first aim was to explore whether there is a significant association between thyroid function and overall cognition in the elderly and whether this association is affected by sex or neurodegenerative status. The second aim was to evaluate whether thyroid function is associated with the odds of being diagnosed with mild cognitive impairment (MCI) at baseline.

## 2. Methods

### 2.1. Study population

Alzheimer’s Disease Neuroimaging Initiative (ADNI) study participants with complete baseline data on demographic characteristics, thyroid function, depression assessment, and overall cognition were included. ADNI, an NIH-funded project (ADNI; National Institutes of Health Grant U19 AG024904 - more in acknowledgments), is a multicenter observational cohort aiming to study the cognitive decline in the elderly and better understand Alzheimer’s disease risks. Dr. Michael Weiner is ADNI’s principal investigator. The first cohort (ADNI 1) started in 2004, and several medical centers around the United States and Canada were involved in recruiting study participants. Following ADNI-1, ADNI-Go, -2, -3, and -4 (ongoing) included more study participants and applied more sophisticated neuropsychiatric and neuroimaging methods. Healthy controls (HC), elderly with MCI, and those with dementia participated in these cohorts. Participants completed written consents for taking part in ADNI. The study was performed according to the Declaration of Helsinki, and ethical approvals were obtained from the local IRBs corresponding to each study center. Study protocols and information on ethical approvals, consent, detailed methods, funding, and data can be found at https://adni.loni.usc.edu.

### 2.2. Thyroid function

Fasting blood samples were collected at baseline. The time between blood collection to freezing did not exceed 120 min. Central thyroid function was explored and thyroid stimulating hormone (TSH) levels, reported in µIU/mL (= mIU/L), were used for this analysis. In the case of repeated measurements, only the first value was retained since the possibility that the study participant received a hormonal supplementation or intervention between the two assessments cannot be precisely verified or excluded. Very low TSH levels (reported as “<0.01”) were converted to 0.01 for the analysis (in three cases). Only participants with TSH < 10 µIU/mL were included, excluding cases with eventual overt hypothyroidism.

Thyroid function was explored in the main analysis based on a continuous variable of TSH (µIU/mL). TSH values 0.4 and 4.5 µIU/mL are clinically relevant cutoff values and are largely considered in literature and guidelines. (22) Clinical cutoff values of “euthyroidism”, “hypothyroidism”, and “hyperthyroidism” depend on various factors and might vary across populations, laboratories, sex groups, during pregnancy, and across age spans. (23) For the sensitivity analysis, thyroid function was classified into three TSH ranges limited by 0.4 and 4.5 µIU/mL, which were considered relevant cutoff values for this study (“TSH < 0.4”, “0.4 ≤ TSH ≤ 4.5”, and “TSH > 4.5”), without further dysthyroidism-related labeling.

### 2.3. Neuropsychological testing

Alzheimer’s Disease Assessment Scale–13-item (ADAS_13_) score was included as a valid tool to assess the overall cognitive status of the ADNI population. Moreover, the Mini-Mental Examination total score (MMSE), Clinical Dementia Rating Scale – Sum of Boxes (CDR-SB), Functional Activity Questionnaire total score (FAQ), and Geriatric Depression Score (GDS) – 15 items total score were added to the descriptive and correlation analyses.

### 2.4. Inclusion criteria

HC and MCI ADNI participants with complete TSH assessment < 10 µIU/mL, complete biometric/demographic (age, sex, educational background, height, and weight) data, complete GDS baseline assessment with GDS total score < 5 (to exclude cases with depression symptoms), (24) and complete ADAS_13_ scores at baseline were eligible. A detailed flow chart is presented in Figure 1.

**Figure 1:**
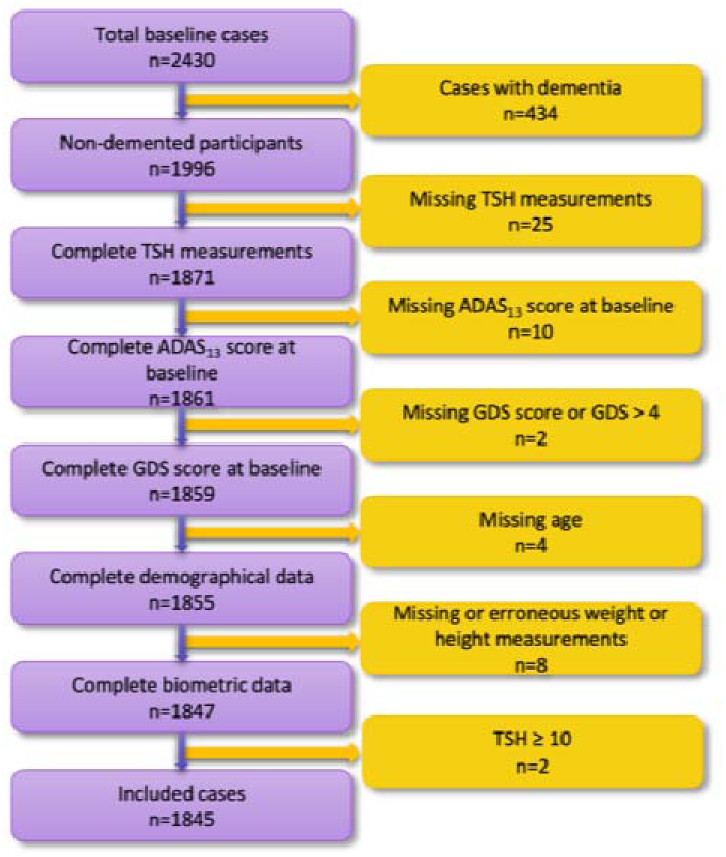
**Flow chart of included cases**

### 2.5. Statistical analysis

The study was based on a cross-sectional analysis of ADNI data at baseline. Median (IQR) and number (%) were reported according to the variable type (continuous and count, respectively). Wilcoxon rank sum test, Pearson’s Chi-squared test, Kruskal-Wallis rank sum test, and Fisher’s exact test were performed depending on the assessed variable and the number of compared groups. The *p*-value was reported for each test. Data was stratified for (1) sex (male and female), and (2) main diagnosis (HC and MCI) as a biomarker of the neurodegenerative state.

Spearman correlation was used to study the association between continuous demographical and cognitive variables, and TSH values. A matrix including the correlation coefficients was visualized in the total population and each stratum.

Linear regression was performed to study the association between TSH levels and ADAS_13_ scores in the total study population, first with a univariable model, then with a multivariable one; adjusted for age (years), sex (male/female), educational background (years of education), racial profile (“White”, “Black”, “Other”), APOE ε4 status (no alleles, one allele, and two alleles), cognition-related main diagnosis (HC or MCI), and BMI (Weight(kg)/Height(m)). ADAS_13_ score was included as a dependent variable and continuous TSH levels (µIU/mL) as an independent variable.

After stratification, univariable linear regression was performed in each stratum. Then, the models were adjusted accordingly for confounding factors:

- **Sex stratum:** age, educational background, racial profile, APOE ε4 status, cognition-related main diagnosis, and BMI.
- **Neurodegeneration-related stratum:** age, sex, educational background, racial profile, APOE ε4 status, and BMI.

Results of linear regressions were reported as regression coefficient (*ß*), 95% confidence interval (*CI*), and *p*-value.

A sensitivity analysis was performed based on TSH ranges as a categorical variable. TSH ranges were included in the linear regression models as independent variables, first in the main study population, and then in the different strata. Both crude and adjusted models, as previously described, were evaluated. The middle TSH range was set as a reference and the regression coefficient *ß*, 95% *CI*, and *p*-value in both groups of “TSH < 0.4” and “TSH > 4.5” were reported in relation to the reference group.

In a second step, a logistic regression analysis was performed to assess the association between thyroid function and the odds of being diagnosed with MCI at baseline. In the main analysis, MCI (binary: yes/no) as a dependent variable, and continuous TSH levels (µIU/mL) as an independent variable were included in the model. First, a crude model was assessed. Then, the same confounders (age, sex, educational background, racial profile, APOE ε4 status, and BMI) were added to the adjusted model. The analysis was repeated in sex strata adjusting accordingly for the confounders (age, educational background, racial profile, APOE ε4 status, and BMI). The sensitivity analysis followed the previously described steps, and TSH ranges were included as predictors in the models. In a further sensitivity analysis, restricted cubic splines were introduced to the model to assess non-linearity in the association. The resulting significant cutoff value was considered to reevaluate the TSH-MCI association, following the strategy of adjustment and stratification already described. The significance level for the null hypothesis was fixed at 0.05 (two-tailed).

Statistical analyses and data visualization were performed with RStudio (version 2024.04.1).

## 3. Results

### 3.1. Characteristics of study participants

A total of 1845 participants with a median age of 73 (IQR: 68, 78) met the inclusion criteria; 958 (52%) of whom were males, 887 were females (48%), 789 (42.76%) were HC, while 1056 (57.24%) had MCI. The median TSH level in the main population at baseline was 1.70 µIU/mL (IQR: 1.15, 2.40), ranged from < 0.01 (three cases that were transformed to 0.01 for the analysis) to 9.58 µIU/mL, and 58 (3.14%) participants had TSH levels < 0.4, 48 (2.60%) TSH > 5, and 1739 (94.25%) had TSH levels between 0.4 and 4.5 µIU/mL. The population characteristics and differences between groups are summarized in Table 1. Further TSH-specific analyses are visualized in Figure 2.

**Figure 2:**
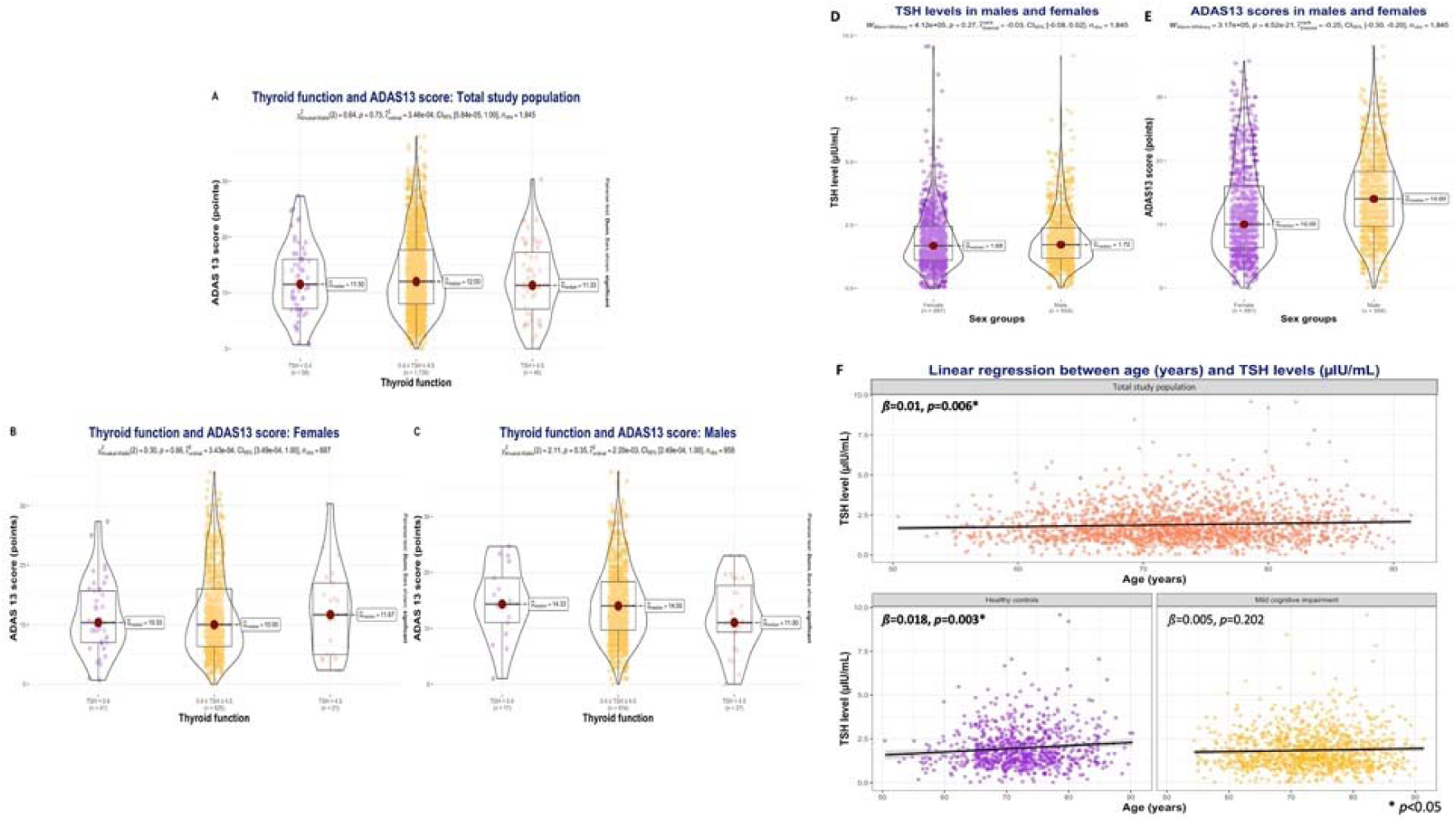
Comparative analyses between study participants and relevant associations. **2. A:** Kruskal-Wallis test of thyroid function and ADAS_13_ score: Total study population. **2. B:** Kruskal-Wallis test of thyroid function and ADAS_13_ score: Females. **2. C:** Kruskal-Wallis test of thyroid function and ADAS_13_ score: Males. **2. D:** Mann-Whitney test of TSH levels in males and females. **2. E:** Mann-Whitney test of ADAS_13_ scores in males and females. **2. F:** Linear regression between age (years) as predicting variable and TSH levels (µIU/mL) as outcome: Total study population, healthy controls, and Mild cognitive impairment

**Table 1:** Characteristics of the total study population and group comparison.

| Characteristic | Total study population |  | Main diagnosis |  |  | Thyroid function |  |  |  |
| --- | --- | --- | --- | --- | --- | --- | --- | --- | --- |
|  | N | Overall<br>N = 1845<sup>1</sup> | Healthy controls<br>N = 789<sup>1</sup> | MCI<br>N = 1056<sup>1</sup> | p-value<sup>2</sup> | TSH < 0.4<br>N = 58<sup>1</sup> | 0.4 ≤ TSH ≤ 4.5<br>N = 1739<sup>1</sup> | TSH > 4.5<br>N = 48<sup>1</sup> | p-value<sup>3</sup> |
| **Age (years)** | 1845 | 73 (68, 78) | 72 (68, 77) | 73 (68, 78) | 0.081 | 71 (67, 78) | 73 (68, 78) | 76 (70, 80) | 0.072 |
| **Sex** | 1845 |  |  |  | **<0.001** |  |  |  | **0.002** |
| Female |  | 887 (48%) | 443 (56%) | 444 (42%) |  | 41 (71%) | 825 (47%) | 21 (44%) |  |
| Male |  | 958 (52%) | 346 (44%) | 612 (58%) |  | 17 (29%) | 914 (53%) | 27 (56%) |  |
| **Marital status** | 1845 |  |  |  | **<0.001** |  |  |  | 0.4 |
| Currently married |  | 1,362 (74%) | 551 (70%) | 811 (77%) |  | 46 (79%) | 1,278 (73%) | 38 (79%) |  |
| Currently not married or unknown |  | 483 (26%) | 238 (30%) | 245 (23%) |  | 12 (21%) | 461 (27%) | 10 (21%) |  |
| **Racial profile** | 1845 |  |  |  | **<0.001** |  |  |  | **0.016** |
| White |  | 1,613 (87%) | 655 (83%) | 958 (91%) |  | 57 (98%) | 1,512 (87%) | 44 (92%) |  |
| Black |  | 148 (8.0%) | 91 (12%) | 57 (5.4%) |  | 1 (1.7%) | 147 (8.5%) | 0 (0%) |  |
| Other |  | 84 (4.6%) | 43 (5.4%) | 41 (3.9%) |  | 0 (0%) | 80 (4.6%) | 4 (8.3%) |  |
| **Educational level (years)** | 1845 | 16.00 (14.00, 18.00) | 16.00 (15.00, 18.00) | 16.00 (14.00, 18.00) | **<0.001** | 16.00 (13.25, 18.00) | 16.00 (14.00, 18.00) | 16.50 (15.75, 18.00) | 0.055 |
| **APOE ε4 status** | 1672 |  |  |  | **<0.001** |  |  |  | >0.9 |
| 0 allele |  | 977 (58%) | 487 (70%) | 490 (50%) |  | 33 (59%) | 916 (58%) | 28 (62%) |  |
| 1 allele |  | 565 (34%) | 190 (27%) | 375 (39%) |  | 19 (34%) | 532 (34%) | 14 (31%) |  |
| 2 alleles |  | 130 (7.8%) | 22 (3.1%) | 108 (11%) |  | 4 (7.1%) | 123 (7.8%) | 3 (6.7%) |  |
| Missing values |  | 173 | 90 | 83 |  | 2 | 168 | 3 |  |
| **ADAS<sub>13</sub> total score** | 1845 | 12 (8, 18) | 8 (5, 12) | 16 (11, 21) | **<0.001** | 12 (7, 16) | 12 (8, 18) | 11 (7, 17) | 0.7 |
| **MMSE total score** | 1845 | 29.00 (27.00, 30.00) | 29.00 (29.00, 30.00) | 28.00 (26.00, 29.00) | **<0.001** | 29.00 (28.00, 29.00) | 29.00 (27.00, 30.00) | 29.00 (27.00, 30.00) | 0.7 |
| **CDR-SB** | 1845 | 0.50 (0.00, 1.50) | 0.00 (0.00, 0.00) | 1.50 (1.00, 2.00) | **<0.001** | 1.00 (0.00, 1.88) | 0.50 (0.00, 1.50) | 0.00 (0.00, 1.00) | **0.030** |
| **FAQ total score** | 1831 | 0.0 (0.0, 2.0) | 0.0 (0.0, 0.0) | 1.0 (0.0, 5.0) | **<0.001** | 1.0 (0.0, 2.8) | 0.0 (0.0, 2.0) | 0.0 (0.0, 0.0) | **0.005** |
| Missing values |  | 14 | 5 | 9 |  | 0 | 14 | 0 |  |
| **BMI** | 1845 | 26.5 (24.1, 29.5) | 26.8 (24.1, 30.0) | 26.3 (24.1, 29.2) | **0.035** | 25.5 (23.2, 29.3) | 26.5 (24.1, 29.5) | 26.4 (24.7, 30.8) | 0.3 |
| **TSH level (μIU/mL)** | 1845 | 1.70 (1.15, 2.40) | 1.74 (1.19, 2.49) | 1.66 (1.09, 2.36) | **0.017** | 0.15 (0.06, 0.26) | 1.71 (1.18, 2.36) | 5.24 (4.80, 6.02) | **<0.001** |
| **Congition-related diagnosis** |  |  |  |  |  |  |  |  | 0.055 |
| Healthy controls |  |  |  |  |  | 18 (31%) | 745 (43%) | 26 (54%) |  |
| MCI |  |  |  |  |  | 40 (69%) | 994 (57%) | 22 (46%) |  |

<sup>1</sup> Median (IQR); n (%), <sup>2</sup> Wilcoxon rank sum test; Pearson's Chi-squared test, <sup>3</sup> Kruskal-Wallis rank sum test; Pearson's Chi-squared test; Fisher's exact test
**ADAS<sub>13</sub>:** Alzheimer's Disease Assessment Scale-13 Items, **APOE:** Apolipoprotein E, **BMI:** Body-Mass Index, **CDR-SB:** Clinical Dementia Rating-Sum of Boxes, **FAQ:** Functional Activities Questionnaire, **MCI:** Mild Cognitive Impairment, **MMSE:** Mini-Mental Status Examination, **TSH:** Thyroid Stimulating Hormone

The median TSH value was significantly lower in the MCI group than in HC (1.66 vs. 1.74, *p*-value: 0.017). The CDR-SB and FAQ scores were higher in those with TSH < 0.4 µIU/mL than the two other TSH range groups (1.00 vs. 0.5 and 0.0, *p*-value=0.030 and 1.00 vs. 0.0 and 0.0, *p*-value=0.005, respectively). No statistically significant differences were observed in the number of MCI cases across the TSH groups.

The correlation analyses are summarized in Figure 3. Only in males, a weak (r_Spearman_= -0.09) but statistically significant correlation (p-value_Spearman_= 0.003) was observed between ADAS_13_ total score and TSH levels.

**Figure 3:**
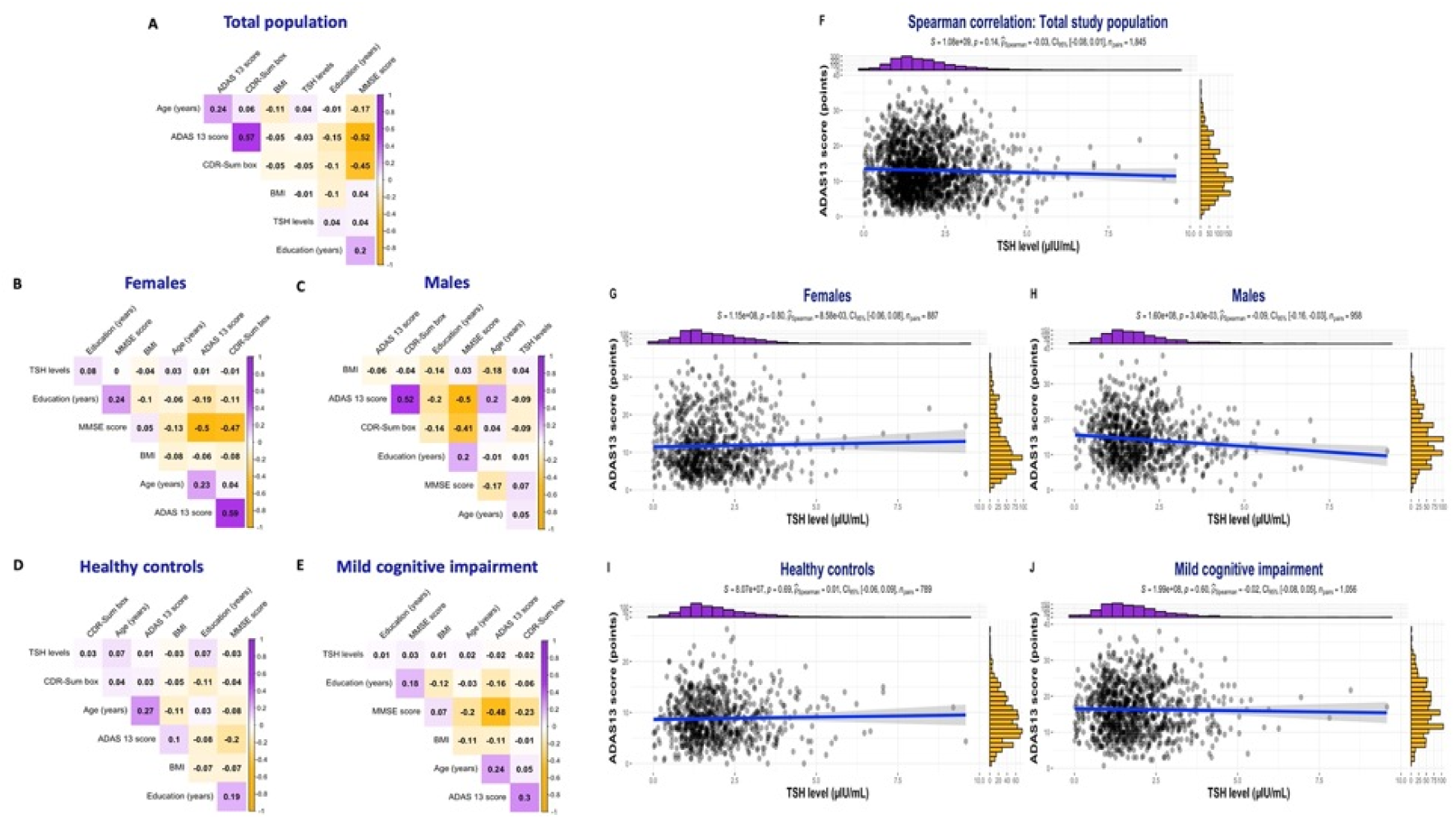
Correlation analyses in the total study population and sex and neurodegeneration-related strata. **3. A:** Correlation matrix: Total study population. **3. B:** Correlation matrix: Females. **3. C:** Correlation matrix: Males. **3. D:** Correlation matrix: Healthy controls. **3. E:** Correlation matrix: Mild cognitive impairment. **3. F:** Spearman correlation between ADAS_13_ total score and TSH level: Total study population. **3. G:** Spearman correlation between ADAS_13_ total score and TSH level: Females. **3. H:** Spearman correlation between ADAS_13_ total score and TSH level: Males. **3. I:** Spearman correlation between ADAS_13_ total score and TSH level: Healthy controls. **3. J:** Spearman correlation between ADAS_13_ total score and TSH level: Mild cognitive impairment.

### 3.2. Thyroid function and overall cognition

The linear regression analysis is detailed in Figure 4. No significant association between TSH levels (µIU/mL) and ADAS_13_ score (points) was found in the main population. In the sex stratum, significant associations were reported only in males, first in the crude model (crude *ß*_Males_: -0.65; 95% *CI*: -1.0, -0.26; *p*-value: 0.001), then after adjustment for confounding factors (adj. *ß*_Males_: -0.40; 95% *CI*: -0.74, -0.07; *p*-value: 0.019).

**Figure 4:**
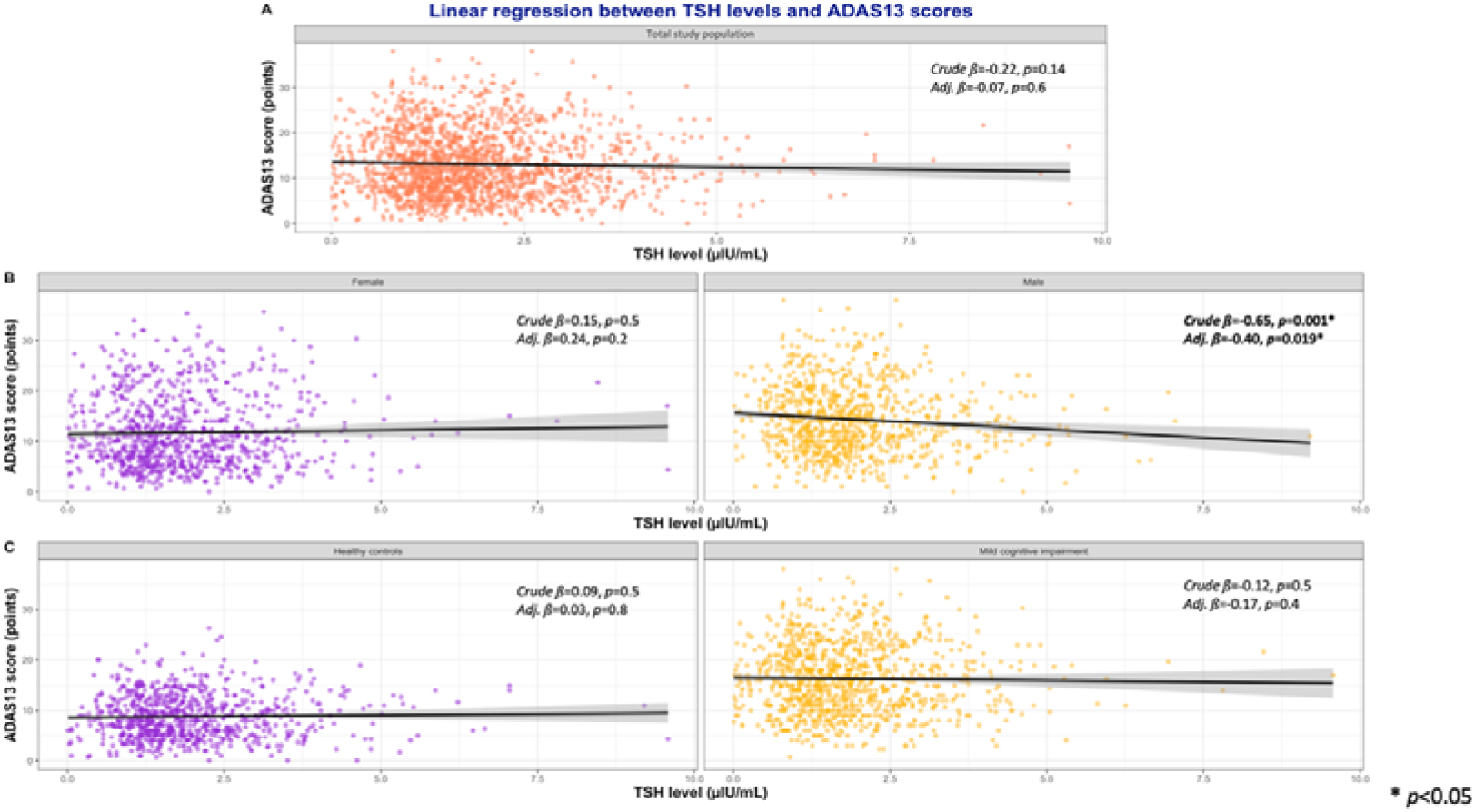
Linear regression analysis of ADAS_13_ total score (points) as an outcome variable and TSH levels (µIU/mL) as a predicting variable in the total study population and sex and neurodegeneration-related strata. **4. A:** Linear regression between TSH levels and ADAS_13_ scores: Total study population. **4. B:** Linear regression between TSH levels and ADAS_13_ scores: Sex strata (“Females” and “Males”). **4. C:** Linear regression between TSH levels and ADAS_13_ scores: Neurodegeneration-related strata (“Healthy controls” and “Mild cognitive impairment”).

Based on TSH ranges, thyroid function was significantly associated with overall cognition only in HC but did not survive the multivariable analysis. Details of the sensitivity analysis are reported in Table 2.

**Table 2:**
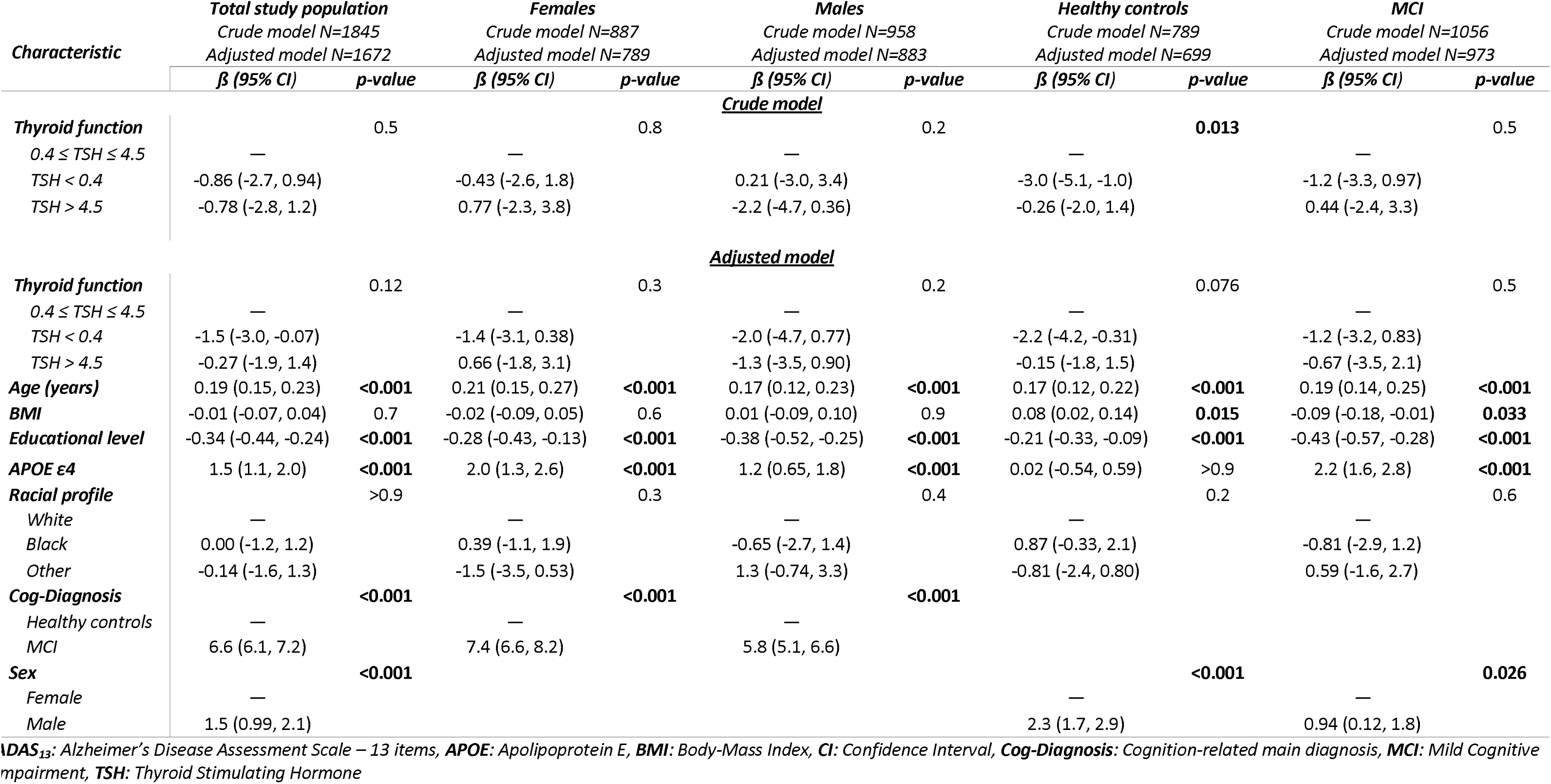
Crude and adjusted models of linear regression between overall cognition (ADAS_13_ total score) as an outcome and thyroid function (TSH ranges) as a predicting variable.

### 3.3. Thyroid function and Mild Cognitive Impairment

The increase in TSH levels of one µIU/mL reduced 10% the odds of being diagnosed with MCI at baseline (crude OR_Total_: 0.90; 95% *CI*: 0.83, 0.98; *p*-value: 0.013) in the total study population. The association remained statistically significant after adjusting for confounders (adj. OR_Total_: 0.87; 95% *CI*: 0.79, 0.95; *p*-value: 0.002). After stratification, only in the male stratum, the increase of TSH levels predicted significantly lower odds of MCI at baseline (crude OR_Males_: 0.82; 95% *CI*: 0.73, 0.93; *p*-value: 0.001), even after adjustment for confounding (adj. OR_Males_: 0.80; 95% *CI*: 0.70, 0.92; *p*-value: 0.001) (Table 3). Sensitivity analysis based on TSH ranges showed similar results with TSH < 0.4 being associated with higher odds of MCI, while TSH > 4.5 with lower odds, compared to the intermediate range group (Table 4).

**Table 3:** Crude and adjusted models of logistic regression for the odds of MCI in the total study population, and in female and male strata – Continuous TSH levels (µIU/mL) as predicting variable.

| Characteristic | Total study population |  |  |  | Females |  |  |  | Males |  |  |  |
| --- | --- | --- | --- | --- | --- | --- | --- | --- | --- | --- | --- | --- |
|  | N | Event N | OR (95% CI) | p-value | N | Event N | OR (95% CI) | p-value | N | Event N | OR (95% CI) | p-value |
| <u>Crude model</u> |  |  |  |  |  |  |  |  |  |  |  |  |
| TSH level ($\mu\text{IU/mL}$) | 1845 | 1056 | 0.90 (0.83, 0.98) | **0.013** | 887 | 444 | 0.97 (0.86, 1.08) | 0.555 | 958 | 612 | 0.82 (0.73, 0.93) | **0.001** |
| <u>Adjusted model</u> |  |  |  |  |  |  |  |  |  |  |  |  |
| TSH level ($\mu\text{IU/mL}$) | 1672 | 973 | 0.87 (0.79, 0.95) | **0.002** | 789 | 401 | 0.94 (0.82, 1.06) | 0.292 | 883 | 572 | 0.80 (0.70, 0.92) | **0.001** |
| Age (years) | 1672 | 973 | 1.00 (0.98, 1.01) | 0.536 | 789 | 401 | 1.00 (0.98, 1.02) | 0.707 | 883 | 572 | 0.99 (0.97, 1.02) | 0.561 |
| BMI | 1672 | 973 | 1.0 (0.97, 1.02) | 0.629 | 789 | 401 | 1.00 (0.97, 1.03) | 0.990 | 883 | 572 | 0.99 (0.95, 1.02) | 0.469 |
| APOE $\epsilon 4$ | 1672 | 973 | 2.07 (1.74, 2.48) | **<0.001** | 789 | 401 | 1.98 (1.56, 2.53) | **<0.001** | 883 | 572 | 2.19 (1.71, 2.85) | **<0.001** |
| Educational level | 1672 | 973 | 0.90 (0.87, 0.94) | **<0.001** | 789 | 401 | 0.91 (0.86, 0.97) | **0.001** | 883 | 572 | 0.89 (0.85, 0.95) | **<0.001** |
| Racial profile | 1672 | 973 |  | **0.024** | 789 | 401 |  | 0.073 | 883 | 572 |  | 0.319 |
| White |  |  | — |  |  |  | — |  |  |  | — |  |
| Black |  |  | 0.53 (0.33, 0.84) |  |  |  | 0.51 (0.28, 0.91) |  |  |  | 0.57 (0.27, 1.27) |  |
| Other |  |  | 1.08 (0.62, 1.88) |  |  |  | 0.90 (0.41, 1.97) |  |  |  | 1.25 (0.58, 2.90) |  |
| Sex | 1672 | 973 |  | **<0.001** |  |  |  |  |  |  |  |  |
| Female |  |  | — |  |  |  |  |  |  |  |  |  |
| Male |  |  | 2.00 (1.62, 2.47) |  |  |  |  |  |  |  |  |  |

APOE: Apolipoprotein E, BMI: Body Mass Index, CI: Confidence Interval, OR: Odds Ratio, TSH: Thyroid Stimulating Hormone

**Table 4:**
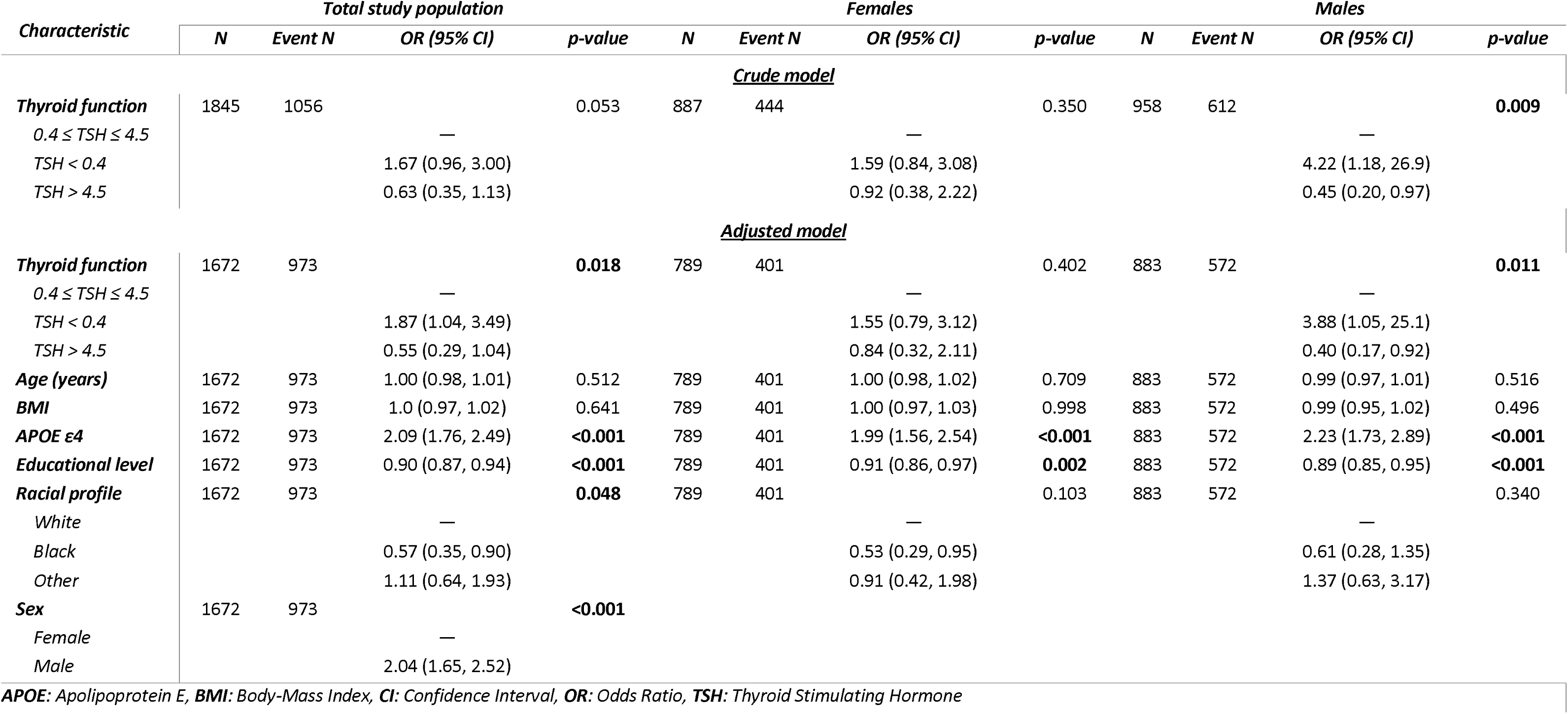
Crude and adjusted models of logistic regression for the odds of MCI in the total study population and in female and male strata – Thyroid function (TSH ranges) as predicting variable.

The introduction of a non-linear factor to the model using the restricted cubic splines, showed that the median TSH value (1.70 µIU/mL) was a significant cutoff value for the association in the total study population and males, but not females (Figure 5). Prediction analysis based on the TSH cutoff value showed that the odds of MCI were significantly higher in people with TSH < 1.70 µIU/mL than in those with TSH levels beyond this cutoff value. The association was significant in the total study population after adjustment for confounders and males, but not females (Table 5). Predicted odds ratios in each stratum in the main and sensitivity analyses are summarized in Figure 6.

**Figure 5:**
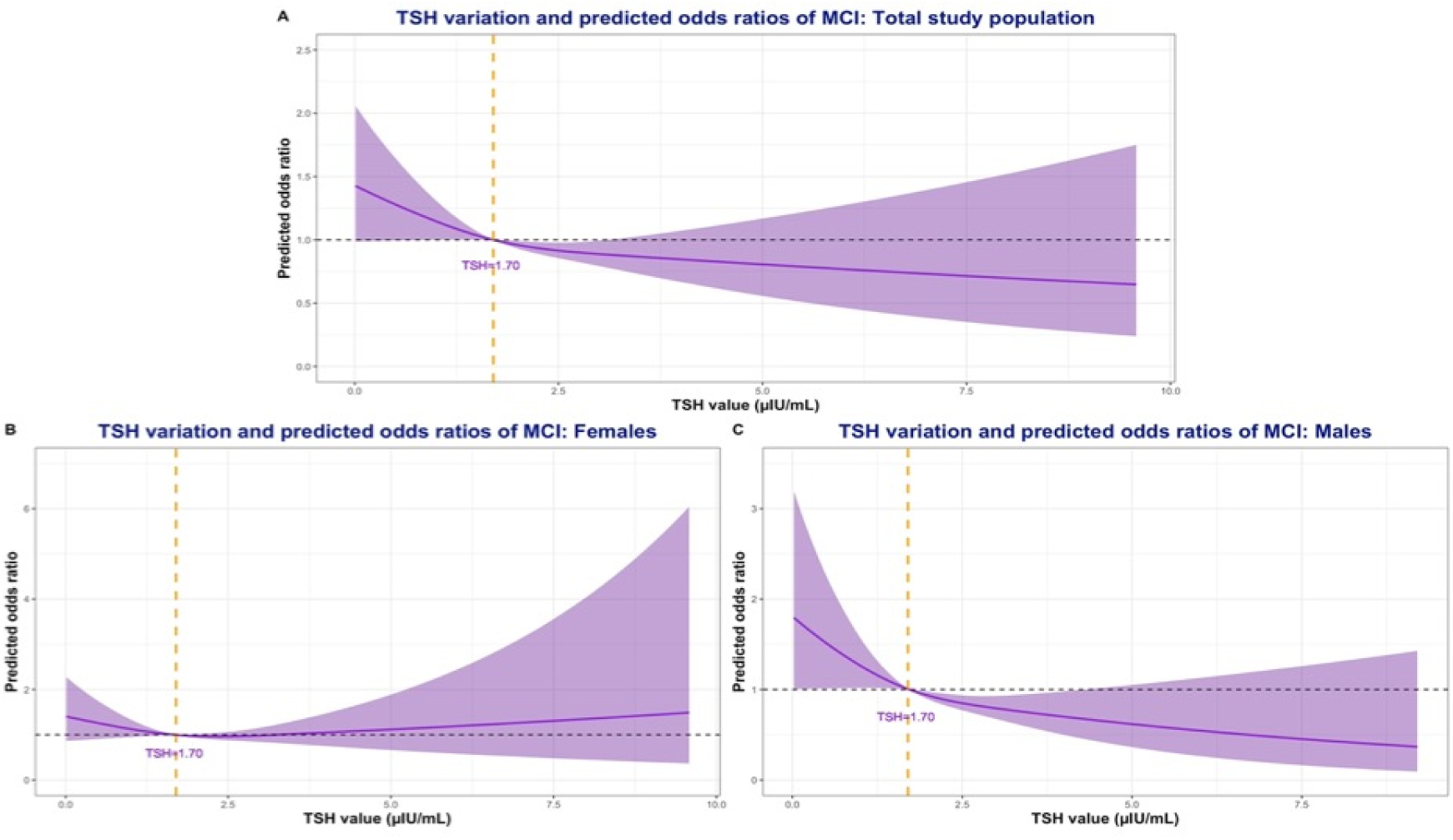
Splines of predicted odds ratios in the total study population and sex strata. **5. A:** TSH variation and predicted odds ratios of MCI: Total study population. **5. B:** TSH variation and predicted odds ratios of MCI: Females. **5. C:** TSH variation and predicted odds ratios of MCI: Males.

**Figure 6:**
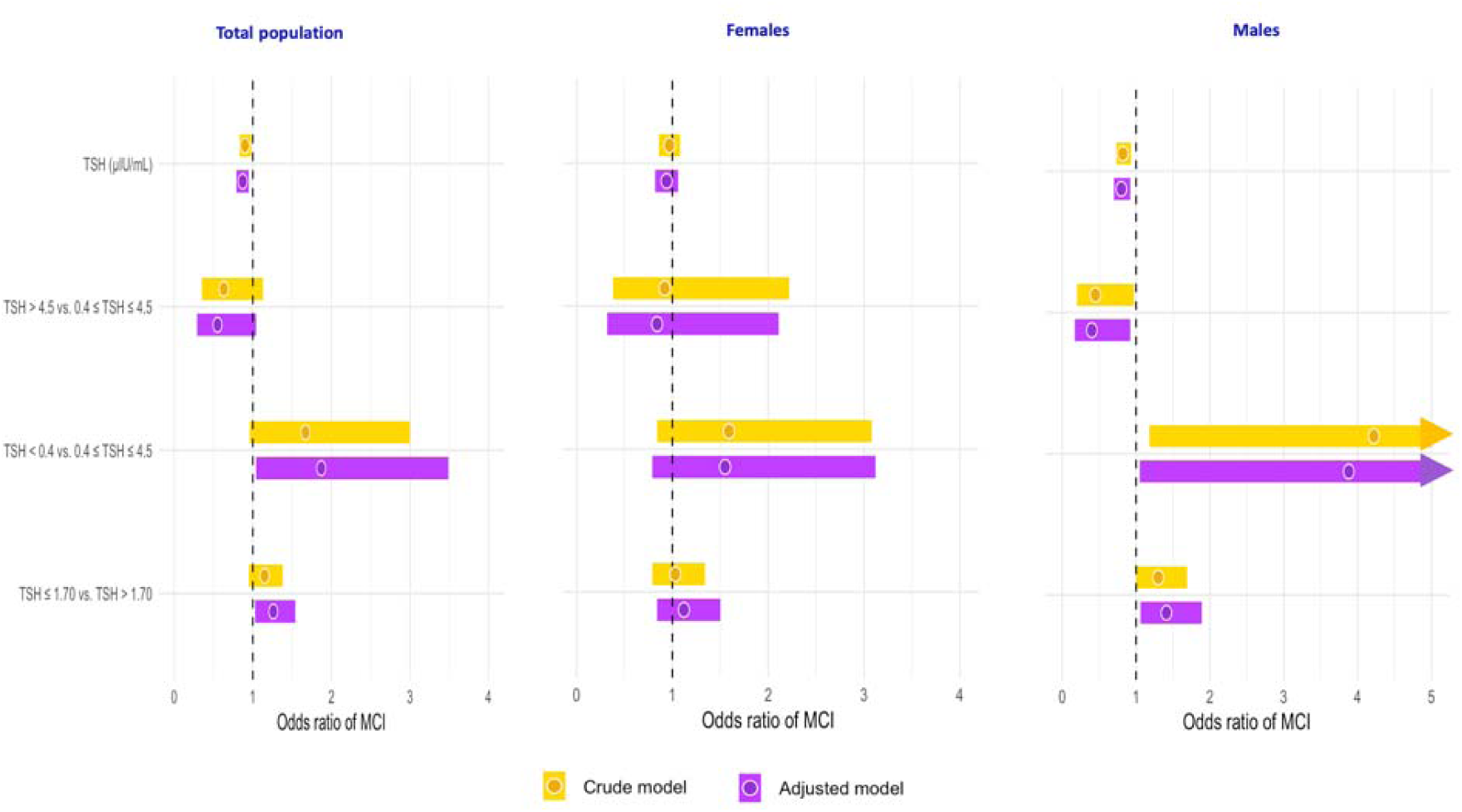
**Summary plots of predicted odds ratios in the total study population and sex strata**

**Table 5:** Crude and adjusted models of logistic regression for the odds of MCI in the total study population and female and male strata – The TSH groups based on a median value as predicting variable.

| *Characteristic* | *Total study population* |  |  |  | *Females* |  |  |  | *Males* |  |  |  |
| --- | --- | --- | --- | --- | --- | --- | --- | --- | --- | --- | --- | --- |
|  | *N* | *Event N* | *OR (95% CI)* | *p-value* | *N* | *Event N* | *OR (95% CI)* | *p-value* | *N* | *Event N* | *OR (95% CI)* | *p-value* |
| **<u>Crude model</u>** |  |  |  |  |  |  |  |  |  |  |  |  |
| ***TSH–median reference*** | 1845 | 1056 |  | 0.147 | 887 | 444 |  | 0.815 | 958 | 612 |  | 0.055 |
| *TSH > 1.70* |  |  | — |  |  |  | — |  |  |  | — |  |
| *TSH ≤ 1.70* |  |  | 1.15 (0.95, 1.38) |  |  |  | 1.03 (0.79, 1.34) |  |  |  | 1.30 (1.00, 1.69) |  |
| **<u>Adjusted model</u>** |  |  |  |  |  |  |  |  |  |  |  |  |
| ***TSH–median reference*** | 1672 | 973 |  | **0.027** | 789 | 401 |  | 0.442 | 883 | 572 |  | **0.019** |
| *TSH > 1.70* |  |  | — |  |  |  | — |  |  |  | — |  |
| *TSH ≤ 1.70* |  |  | 1.26 (1.03, 1.54) |  |  |  | 1.12 (0.84, 1.50) |  |  |  | 1.41 (1.06, 1.89) |  |
| ***Age (years)*** | 1672 | 973 | 1.0 (0.98, 1.01) | 0.488 | 789 | 401 | 1.00 (0.97, 1.02) | 0.667 | 883 | 572 | 0.99 (0.97, 1.02) | 0.566 |
| ***BMI*** | 1672 | 973 | 1.0 (0.97, 1.02) | 0.644 | 789 | 401 | 1.00 (0.97, 1.03) | 0.998 | 883 | 572 | 0.99 (0.95, 1.02) | 0.481 |
| ***APOE ε4*** | 1672 | 973 | 2.08 (1.75, 2.48) | **<0.001** | 789 | 401 | 1.98 (1.56, 2.53) | **<0.001** | 883 | 572 | 2.20 (1.71, 2.85) | **<0.001** |
| ***Educational level*** | 1672 | 973 | 0.90 (0.87, 0.94) | **<0.001** | 789 | 401 | 0.91 (0.86, 0.97) | **0.001** | 883 | 572 | 0.89 (0.84, 0.94) | **<0.001** |
| ***Racial profile*** | 1672 | 973 |  | **0.032** | 789 | 401 |  | 0.080 | 883 | 572 |  | 0.354 |
| *White* |  |  | — |  |  |  | — |  |  |  | — |  |
| *Black* |  |  | 0.54 (0.34, 0.86) |  |  |  | 0.52 (0.28, 0.92) |  |  |  | 0.59 (0.27, 1.31) |  |
| *Other* |  |  | 1.07 (0.62, 1.86) |  |  |  | 0.89 (0.41, 1.95) |  |  |  | 1.26 (0.58, 2.90) |  |
| ***Sex*** | 1672 | 973 |  | **<0.001** |  |  |  |  |  |  |  |  |
| *Female* |  |  | — |  |  |  |  |  |  |  |  |  |
| *Male* |  |  | 1.99 (1.61, 2.46) |  |  |  |  |  |  |  |  |  |

*APOE*: Apolipoprotein E, *BMI*: Body-Mass Index, *CI*: Confidence Interval, *OR*: Odds Ratio, *TSH*: Thyroid Stimulating Hormone

## 4. Discussion

The high number of study participants, the exclusion of dementia, overt hypothyroidism, and depression as relevant confounders, and the sex stratification were major strengths of this study. The first outcome was the significant association between TSH levels and ADAS_13_ scores. Although no statistically significant results have been found in the main study population, significant associations between thyroid function and overall cognition were revealed in male participants after stratifying for sex. The second outcome was the value of TSH in significantly predicting MCI diagnosis at baseline, particularly in elderly males. The association between TSH and MCI diagnosis was particularly significant in those with TSH levels lower than the median TSH value (1.70 µIU/mL).

### 4.1. Thyroid function and cognition

The association between TSH levels and cognition in the elderly was controversial in the literature. While some studies on older euthyroid adults did not identify significant associations between TSH and free T4 (FT4, thyroxine) levels and cognition, (25) other studies reported significant associations between lower thyroid hormones – free T3 (FT3, tri-iodothyronine) and cognitive impairment. (26)

In studies where both euthyroid patients as well as patients with subclinical hypo- and hyperthyroidism were included, no significant association was found between thyroid hormones and MMSE scores, despite the higher age of included patients (85 years). (27) TSH did not show a significant association with CERAD subdomains in patients of 75 years and older, but higher TSH levels were significantly correlated with better CDR scores than lower levels. (28)

While those studies focused on associations between thyroid function and cognitive tests, a further prospective cohort did not show a significant value of TSH in predicting the progression from MCI to dementia within three years in euthyroid and hypothyroid elderly patients. (29) In contrast, lower TSH levels at baseline predicted progression to MCI or dementia over five years in Korean euthyroid and at baseline non-demented elderly. (30) The sample size and inclusion criteria seem to affect the reported results and explain the controversies in published data. The current study was based on estimating the overall cognition using ADAS_13_ scores, but the outcome of published data was sometimes based on continuous scores (MMSE, CERAD), or binary outcomes such as conversion to MCI or dementia, and particular interest was given to exploring the associations in higher TSH levels or hypothyroidism.

TSH levels tend to increase with age, and physiological ranges have to be adapted accordingly. Subclinical hypothyroidism is a more common condition in the elderly and does not necessarily implicate a pathological status or necessitate hormone therapy. (17, 18) In the current study, higher age was associated with higher TSH levels in the total study population and after stratification in healthy controls but not in participants with MCI. In a placebo-controlled randomized clinical trial, hormonal supplementation in older adults with subclinical hypothyroidism did not show a significant impact on cognitive function. (31) In older populations (85 years), a longitudinal decrease in TSH levels was associated with a concomitant reduction in MMSE scores over a three- and five-year follow-up period. (32) Thyrotoxicosis is a rarer condition in which patients present very high thyroid hormones either of endogenous or exogenous etiology (corresponding to very low TSH). In a study including 65931 elderly patients aged 65 years and older, thyrotoxicosis was associated with a significantly higher risk of cognitive decline. (33) Among the included cases from the ADNI cohort, three participants had TSH levels of < 0.01 and were converted to 0.01 for the analysis. Despite that, the present study confirmed that lower TSH levels (independently from thyrotoxicosis) were associated with worse cognitive scores and a higher risk of being diagnosed with MCI. These results were particularly significant under a cutoff value corresponding to the median TSH value in males, but not females.

### 4.2. Effect of sex on the relationship between thyroid function and cognition

Samuels and colleagues studied in a cross-sectional and longitudinal design eventual associations between the variation of thyroid function within age-adapted normal ranges, and cognition in older men (≥ 65 years). Neither TSH nor FT_4_ showed statistically significant results. (34) This contradicts another study in elderly men aged between 70 and 89 years, where higher FT_4_ (but not TSH) levels were associated with higher dementia incidence. (35) In women between 30 and 64 years old, higher TSH levels were significantly associated with a faster decline in clock-command scores. (36) Moreover, in a cross-sectional analysis of 2563 euthyroid elderly aged between 50 and 80 years, higher TSH levels were significantly associated with the diagnosis of MCI only in women. (37)

The paradoxical association between thyroid function and cognition seen in male (negative association) and female (positive association) strata was an interesting outcome in the current study. However, the statistical significance of the analysis was only observed in elderly males. The present findings confirm previous studies in favor of a sex-dependent association between thyroid hormones and cognition in the elderly. (38) Yet, eventual sex-specific confounding factors need to be better understood.

### 4.3. Effect of age on the relationship between thyroid function and cognition

Thyroid function affects the elderly differently across ages. While studies in the elderly reported controversial results, a large cross-sectional analysis including 10362 middle-aged adults (mean: 49.5 ±7.4 years) showed that lower TSH levels are associated with impaired executive function. (39) Moreover, higher TSH levels were associated with poorer cognition in adults under 59 years and better performance in those older than 60 years. (40)

### 4.4. Effect of neurodegeneration on thyroid-cognition relationship

Although neither elderly with dementia nor TSH measurements in cerebrospinal fluid were included in the current analysis, studies have reported a biological association between thyroid hormones and Alzheimer’s pathology, even within euthyroid elderly. (41–43) While stratifying for the biomarker of the neurodegenerative status (HC vs. MCI) did not show significant associations between thyroid function and cognition after adjusting for confounding factors, the association between TSH and Alzheimer’s disease biological biomarkers was out of the scope of this study. Further investigations are needed, particularly to assess whether sex impacts similarly this association.

### 4.5. Limitations

The first limitation of our study is related to the lower number of participants with thyroid levels beyond cutoff values compared to participants with relatively “normal” TSH levels at baseline. This might be related to the healthcare awareness and compliance in persons with health-seeking behavior, who are more likely to consent to be study participants in a longitudinal cohort like ADNI. Moreover, people with severe comorbidities were not eligible for the study. Furthermore, the study was only based on TSH levels without evaluating the potential effect of peripheral thyroid hormones (FT_3_ and FT_4_). This is mainly due to the lack of additional laboratory data in the accessible ADNI database.

The second limitation is the non-inclusion of the medical history related to thyroid pathologies, particularly whether there was an underlying supplemented hypothyroidism or an eventual history of thyroid (surgery/radioiodine) ablation. Moreover, information on potential concomitant autoimmune diseases such as Graves or Hashimoto thyroiditis was out of the scope of the study. This is primarily due to the coding system’s reliability, based on self-reporting of medical history. This might consequently lead to higher risks of recall or assessment bias. The main exposure of the study was the central thyroid function based on TSH levels independently from etiologies and endocrine comorbidities.

Larger population-based longitudinal studies are needed to infer a potential causal effect and assess the associations in risk groups, particularly to better understand the role of sex in modulating the observed effects.

### 4.6. Perspectives and significance

This study highlighted the importance of sex in modulating the association between thyroid function and cognition. Low TSH levels are associated with higher odds of being diagnosed with MCI, and this association might be revealed only after stratifying for sex and exploring separately males and females. TSH-related cognitive impairment is largely believed to be a reversible condition, especially when associated with overt hypothyroidism. Thus, the reversibility of cognitive decline associated with lower TSH levels has to be tested in further randomized studies. It is also important to replicate the findings in different populations and test for non-linearity. The predictive value and the biological significance of the median TSH level in older males need to be more studied and better understood. The sex-dependent neurobiology of cognition in the context of thyroid function needs to be further explored to decrypt the paradoxical differences between males and females.

## 5. Conclusions

This study provides additional arguments supporting the theory of a multifaceted brain-thyroid interaction in advanced ages. Sex infers significantly the association between thyroid function and cognition in older populations, and the association is subject to a cutoff value. Lower TSH values were associated with cognitive decline in elderly males but not females, but the underlying mechanisms need to motivate further research.

## Data Availability

All data produced are available online at https://adni.loni.usc.edu

## Declarations

### Ethics approval and consent to participate

Ethical approvals were obtained from corresponding IRBs in each recruitment site. Written consent was obtained from every ADNI study participant. More information is available at http://adni.loni.usc.edu

### Consent for publication

Not applicable. Brain and Thyroid vectors used in the graphical abstract are designed by Freepik (License free).

### Availability of data and materials

The datasets supporting the conclusions of this article are available at http://adni.loni.usc.edu

### Author’s contributions

AH has full access to all of the data and takes responsibility for the integrity of the data and the accuracy of the analysis, visualization, drafting, and editing of the manuscript.

## Acknowledgments

“Data collection and sharing for the Alzheimer’s Disease Neuroimaging Initiative (ADNI) is funded by the National Institute on Aging (National Institutes of Health Grant U19 AG024904). The grantee organization is the Northern California Institute for Research and Education. In the past, ADNI has also received funding from the National Institute of Biomedical Imaging and Bioengineering, the Canadian Institutes of Health Research, and private sector contributions through the Foundation for the National Institutes of Health (FNIH) including generous contributions from the following: AbbVie, Alzheimer’s Association; Alzheimer’s Drug Discovery Foundation; Araclon Biotech; BioClinica, Inc.; Biogen; Bristol-Myers Squibb Company; CereSpir, Inc.; Cogstate; Eisai Inc.; Elan Pharmaceuticals, Inc.; Eli Lilly and Company; EuroImmun; F. Hoffmann-La Roche Ltd and its affiliated company Genentech, Inc.; Fujirebio; GE Healthcare; IXICO Ltd.; Janssen Alzheimer Immunotherapy Research & Development, LLC.; Johnson & Johnson Pharmaceutical Research &Development LLC.; Lumosity; Lundbeck; Merck & Co., Inc.; Meso Scale Diagnostics, LLC.; NeuroRx Research; Neurotrack Technologies; Novartis Pharmaceuticals Corporation; Pfizer Inc.; Piramal Imaging; Servier; Takeda Pharmaceutical Company; and Transition Therapeutics.”

